# Incidence of SARS-CoV-2 infection and associated risk factors among staff and residents at homeless shelters in King County, Washington: an active surveillance study

**DOI:** 10.1101/2023.05.25.23290471

**Authors:** Julia H. Rogers, Sarah N. Cox, Amy C. Link, Gift Nwanne, Peter D. Han, Brian Pfau, Eric J. Chow, Caitlin R. Wolf, Michael Boeckh, James P. Hughes, Elizabeth Halloran, Timothy M. Uyeki, Mia Shim, Jeffrey Duchin, Janet A. Englund, Emily Mosites, Melissa A. Rolfes, Lea A. Starita, Helen Y. Chu

**Affiliations:** Division of Allergy and Infectious Diseases, Department of Medicine, University of Washington, Seattle, Washington, USA; Department of Epidemiology, University of Washington, Seattle, Washington, USA; Vaccine and Infectious Disease Division, Fred Hutchinson Cancer Center, Seattle, Washington, USA; Department of Global Health, University of Washington, Seattle, Washington, USA; Brotman Baty Institute for Precision Medicine, Seattle, Washington, USA; Department of Biostatistics, University of Washington, Seattle, Washington, USA; Influenza Division, National Center for Immunization and Respiratory Diseases, Centers for Disease Control and Prevention, Atlanta, Georgia, USA; Public Health – Seattle & King County, Seattle, Washington, USA; Department of Medicine, University of Washington, Seattle, Washington, USA; Division of Pediatric Infectious Diseases, Department of Pediatrics, University of Washington, Seattle Children’s Research Institute, Seattle, Washington; Office of the Deputy Director for Infectious Diseases, Centers for Disease Control and Prevention, Atlanta, Georgia, USA; Virology Division, Department of Laboratory Medicine and Pathology, University of Washington, Seattle, Washington, USA

## Abstract

Homeless shelter residents and staff may be at higher risk of SARS-CoV-2 infection. However, SARS-CoV-2 infection estimates in this population have been reliant on cross-sectional or outbreak investigation data. We conducted routine surveillance and outbreak testing in 23 homeless shelters in King County, Washington to estimate the occurrence of laboratory-confirmed SARS-CoV-2 infection and risk factors during 1/1/2020-5/31/2021. Symptom surveys and nasal swabs were collected for SARS-CoV-2 testing by RT-PCR for residents aged ≥3 months and staff. We collected 12,915 specimens from 2,930 unique participants. We identified 4.74 (95% CI 4.00 – 5.58) SARS-CoV-2 infections per 100 individuals (residents: 4.96, 95% CI 4.12 – 5.91; staff: 3.86, 95% CI 2.43 – 5.79). Most infections were asymptomatic at time of detection (74%) and detected during routine surveillance (73%). Outbreak testing yielded higher test positivity compared to routine surveillance (2.7% vs. 0.9%). Among those infected, residents were less likely to report symptoms than staff. Participants who were vaccinated against seasonal influenza and were current smokers had lower odds of having an infection detected. Active surveillance that includes SARS-CoV-2 testing of all persons is essential in ascertaining the true burden of SARS-CoV-2 infections among residents and staff of congregate settings.

## Background

The coronavirus disease 2019 (COVID-19) pandemic has posed unprecedented challenges to the more than 580,000 people experiencing homelessness (PEH) estimated in the U.S. on a single night in 2020.(1) These challenges exacerbated systemic inequities that adversely impact existing health conditions, access to health care, and work and living conditions, potentiating a disproportionate risk of severe acute respiratory syndrome coronavirus 2 (SARS-CoV-2) infection and its subsequent clinical manifestations among PEH.(2) Difficulty with social distancing and high prevalence of chronic diseases led to early concern that PEH in shelters would be at greater risk of COVID-19 complications.(3,4) Homeless service providers may also face greater risk of exposure to SARS-CoV-2 as a result of working in a congregate living setting.(5) Because of this concern, many communities worked together with homeless service providers to put protective measures in place for residents.(6)

However, implementation of consistently available SARS-CoV-2 testing in shelters has been challenging.(7) Most studies have relied on cross-sectional data or been centered on single outbreak investigations in specific geographies, limiting their generalizability.(5,8–10) Robust testing data are vital to mitigating viral transmission through early identification and isolation of cases.(11)

King County, Washington has one of the largest populations of PEH in the U.S (11,751 on a single night in 2020).(12) We previously described early characteristics of SARS-CoV-2 in King County shelters and detected a 2% test positivity rate.(9) Important questions remain as to whether certain individual or shelter-level characteristics are associated with higher risk of infection among shelter residents and staff. In this study, we aimed to characterize the burden of disease among a diverse shelter population using data collected from active surveillance. We captured temporal trends and estimated the incidence and associated risk factors of SARS-CoV-2 infection among shelter residents and staff.

## Methods

### Study design overview and population

We conducted an active community-based surveillance study of SARS-CoV-2 cases in shelters across Seattle-King County from 1/1/2020-5/31/2021. This was a sub-study of a multiyear, cluster randomized trial (CRT) of onsite testing and treatment for influenza at nine shelters that took place from 10/1/19-5/31/21 (registration number: NCT04141917).(13) From 1/1/2020-3/31/2020 eligibility for participation included: aged ≥3 months, residents at a shelter study site, and having cough alone or ≥2 new or worsening acute respiratory illness (ARI) symptoms with onset in the past 7 days. Once a month, eligibility was extended to residents aged ≥3 months regardless of symptoms. In response to the identification of SARS-CoV-2 community transmission in Washington State on 2/24/2020,(14) the first year of the influenza trial intervention was paused on 4/1/2020 and eligibility was expanded to include all shelter staff and residents aged ≥3 months, regardless of symptoms (Figure S1). Participants eligible for COVID-19 testing did not have to be eligible for the influenza test-and-treat intervention during the study period, but they could elect to take part during surveillance months when the intervention was available for those that met additional criteria.(15)

### Study setting and sampling strategy

Participants were recruited three to six days per week by research staff at selected shelters using two mechanisms: routine surveillance and outbreak testing events. These mechanisms have been previously described(9); in brief, routine surveillance involved self-selected participation at staffed kiosks in shelters during standardized days and times. COVID-19 outbreak testing was initiated on 3/30/2020 (and conducted intermittently thereafter) in collaboration with Public Health Seattle-King County (PHSKC) with single day intensive testing for all available residents and staff at shelters where ≥ 1 SARS-CoV-2 infections were recently detected. Individual participants were not followed longitudinally, but eligible individuals may have multiple encounters throughout the study period as routine testing was used as a study recruitment tool and proactive public health strategy. Study participation was limited to once weekly, unless new or worsening ARI symptoms developed, in which case an individual could re-enroll within seven days. Participants were recruited from 23 shelters total over the study period; routine surveillance and outbreak testing were conducted at 15 shelters, while outbreak-only testing was conducted at the other 8 shelters. Routine surveillance was conducted concurrently at 9 shelters at any given time over the study period; six of these shelters relocated staff and residents to new facilities to enable improved adherence to COVID-19 infection and prevention control measures, resulting in 15 shelters total where routine surveillance occurred. Research activities were immediately initiated following these relocations (Table S1).

### Measures

The primary outcome was incidence of reverse transcription polymerase chain reaction (RT-PCR)-confirmed SARS-CoV-2 infection, 1/1/2020-5/31/2021. All inconclusive testing results were classified as SARS-CoV-2 infections per PHSKC and Washington Department of Health guidelines.(16) Incidence is customarily defined as either the proportion of a population at risk that develops the outcome of interest over a specified time period (cumulative incidence) or the count of incidence cases divided by the aggregate amount of at-risk experience (incidence rate). This study describes incident infections detected through repeated cross-sectional testing in an open population of individuals that experienced homelessness or worked at a shelter at some point in study period but were not necessarily at risk for its entirety; we were not able to capture individual time-at-risk.

Survey data were collected electronically on a tablet at time of nasal swab collection from residents and staff. Data included participant sex, date of birth (DOB), race, ethnicity (Hispanic or Latino vs. non-Hispanic or Latino), self-reported current season receipt of an influenza vaccine, underlying medical conditions, status as shelter staff versus resident, highest education level obtained, health insurance status, employment status, and self-reported receipt of any COVID-19 vaccine doses. Smoking status included any current use of tobacco products, e-cigarettes, or vape pens. Underlying conditions included asthma, blood disorders, cancer, chronic obstructive pulmonary disease or emphysema, chronic bronchitis, immunosuppression, liver disease, heart disease, diabetes, neurologic conditions, or aspirin therapy. All survey data characteristics presented in this analysis were collected from both residents and staff, with the exception of sleeping arrangements and duration of homelessness. Sleeping arrangements were reported only by shelter residents and categorized as communal, open-plan cubicles, or private room/shared family room. Communal included sleeping in a congregate space with bunk beds, bed mats, or rooms shared with more than one family. Enrollments per unique participant was defined as the number of survey responses collected from the same participant over the study period. All variables were determined by self-report.

Participant encounters with one or more new or worsening ARI symptoms with onset in the past seven days were defined as symptomatic, and those without any new or worsening symptoms in the past seven days were defined as asymptomatic. This phrasing aimed to specifically distinguish acute symptoms indicative of respiratory viral infection in a population with high rates of chronic illness.(17) Participants with ARI symptoms also had symptom duration data collected in response to the question, “*When did the symptoms you mentioned in the beginning of this survey become new or worsening?*” Influenza-like illness (ILI) was defined as having a fever and either cough or sore throat. COVID-19–like illness (CLI) was defined as fever and cough or fever and increased difficulty breathing.

### Specimen Collection

Mid-turbinate nasal swabs were obtained using a sterile nylon flocked nasal swab (Copan Diagnostics) by a member of the research staff until 3/6/2020, after which participants self-collected a mid-turbinate nasal swab while observed by study staff. Due to supply shortages, anterior nare swabs replaced the use of mid-turbinate swabs from July 2020 through October 2020. See “Supplementary Material” for specimen testing details.

### Statistical Analysis

The primary unit of analysis was unique participants, with corresponding individual-level characteristics taken from their last survey response. Participant encounters from unique individuals were dropped in this analysis following a positive or inconclusive test result; no persistent-positive test results or repeat infections were included in the analysis (n=543; Figure S2).

Incidence of SARS-CoV-2 infection was expressed as cases per 100 unique participants at risk, and described by age group, sex, race, ethnicity, and shelter type. The overall incidence of SARS-CoV-2 infection was calculated by dividing the total number of confirmed cases across all shelters by the total number of unique participants tested over the study period with 95% confidence intervals (CI).

Temporal trends of SARS-CoV-2 test positivity were also reported by epidemiologic week, a standardized method of counting weeks to allow for data comparison year after year. (18) For the specific purpose of calculating and depicting temporal trends of SARS-CoV-2 test positivity, we included all tests collected as the unit of analysis, regardless of participants’ frequency of testing.

Participant-level characteristics were summarized by using frequencies, percentages, and interquartile ranges. We used a chi-square test for independence for categorical variables (or Fisher’s exact test for when cells had less than 10 observations) and a t-test for continuous variables of individual-level participant characteristics and SARS-CoV-2 infection, separately among shelter residents and shelter staff. To estimate corresponding adjusted associations with SARS-CoV-2 infection among staff and residents separately (Table 3a) and symptomatic COVID-19 disease among all infected participants (Table 3b), respectively, we used generalized linear mixed models (GLMM), treating shelter as a random effect. Variables were selected for the models presented in Table 3a and Table 3b using a causal diagram approach. Risk factors included in the final multivariable models were checked for multicollinearity and convergence issues due to excessive missingness (e.g., “duration of homelessness” which was not asked of shelter staff). Descriptive statistics for test-level variables (Table 1c) are presented but were not considered for inclusion in the multivariable models as we were primarily interested in fixed individual-level exposures.

**Table 1a.**
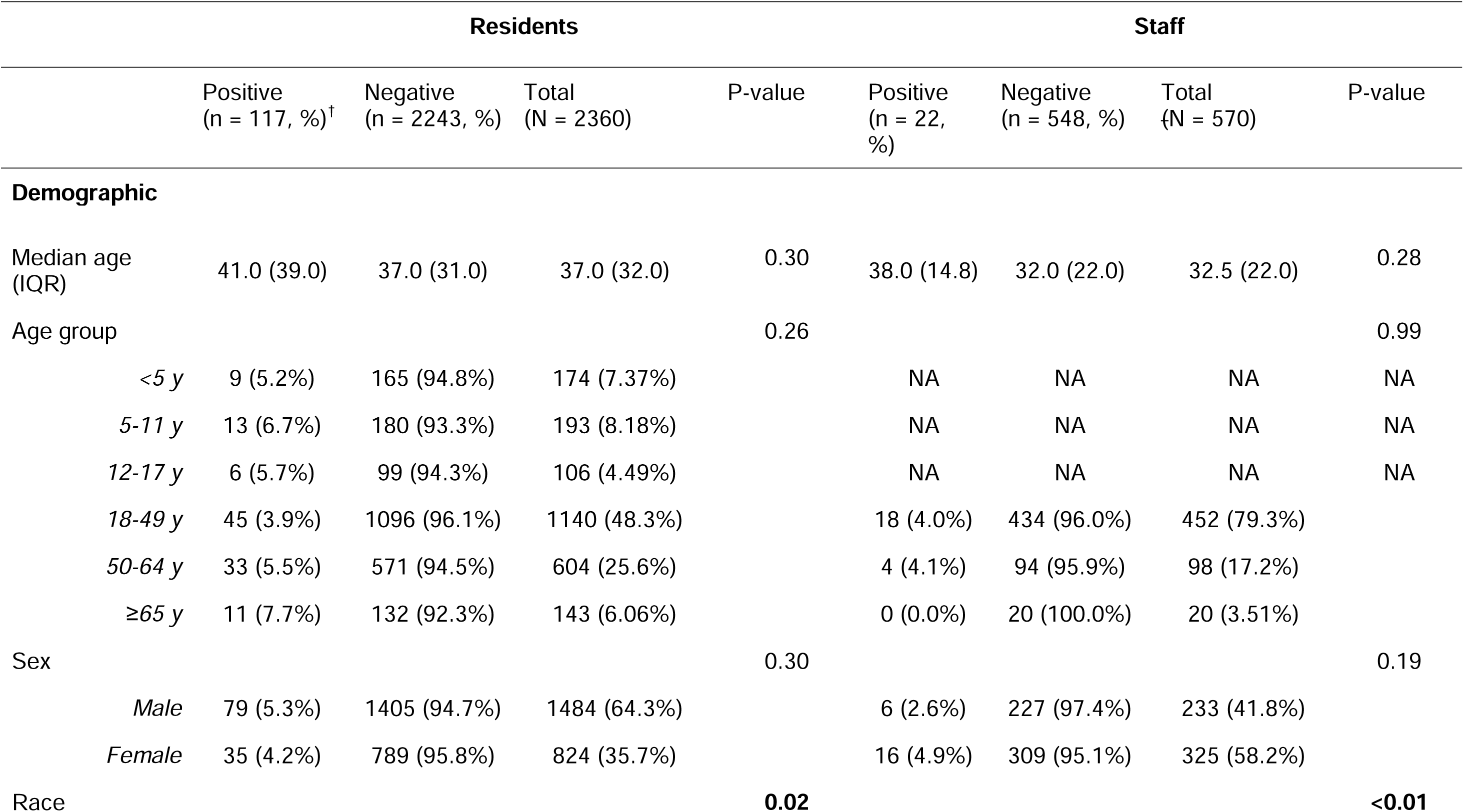
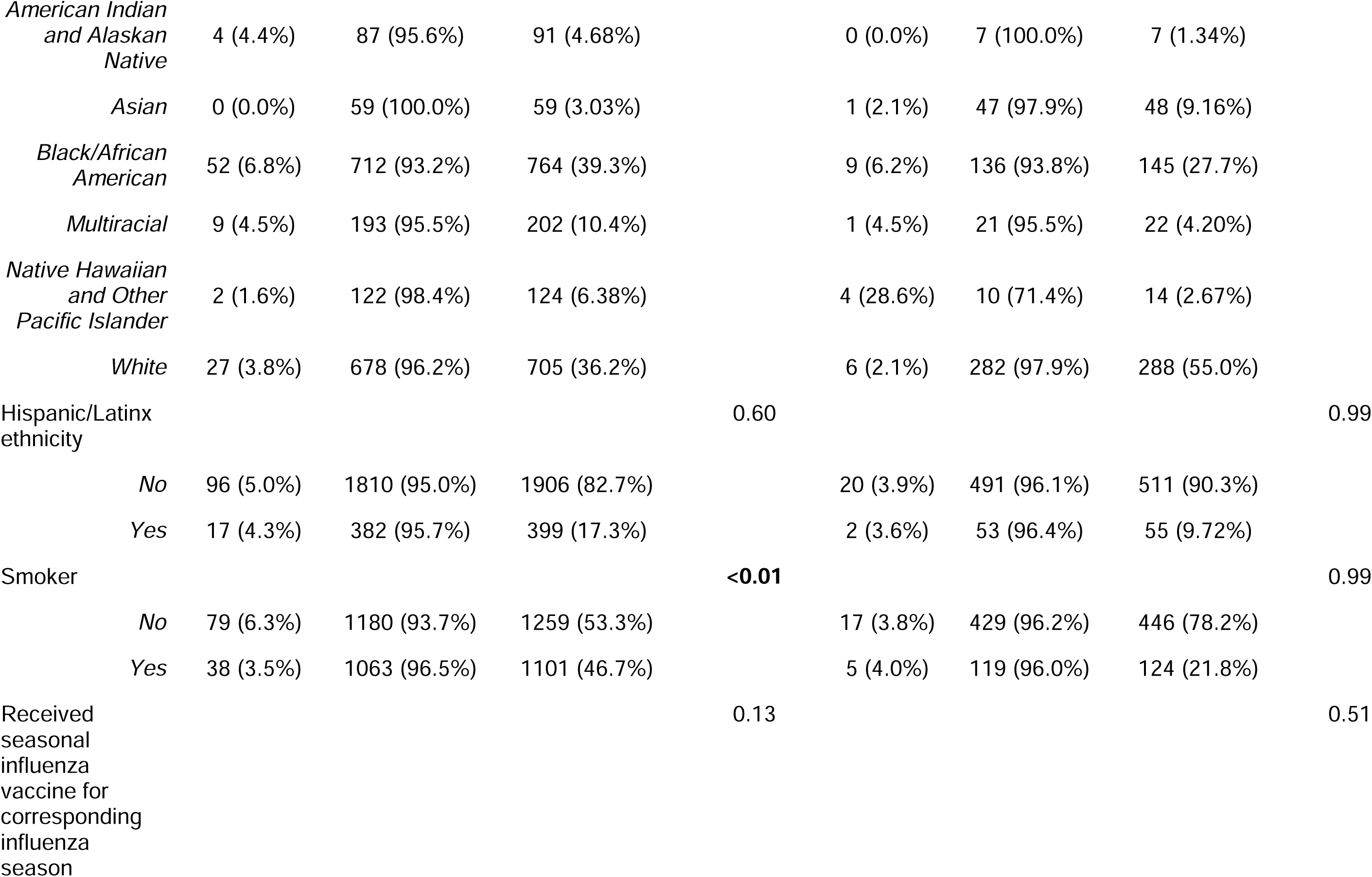
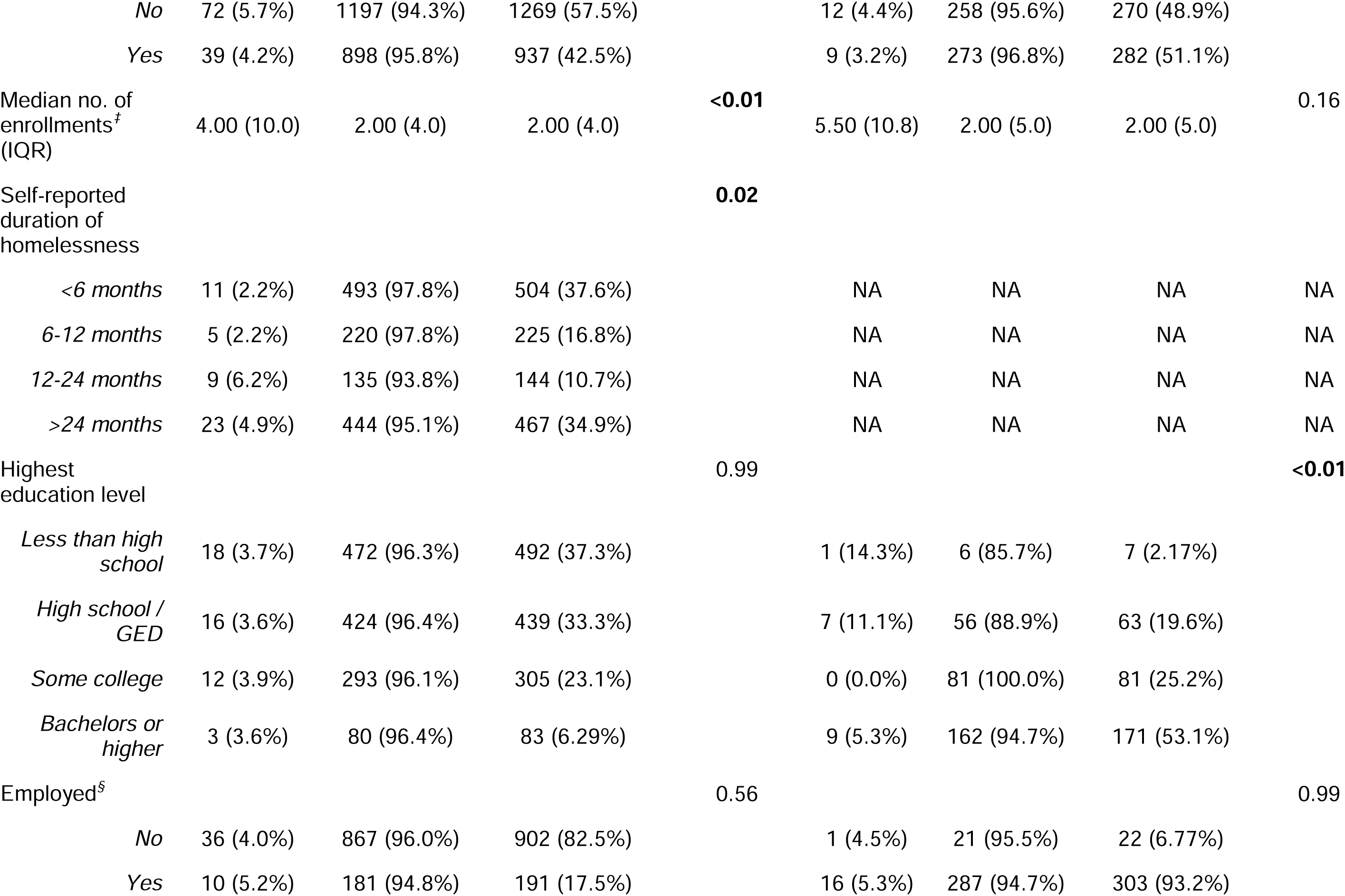
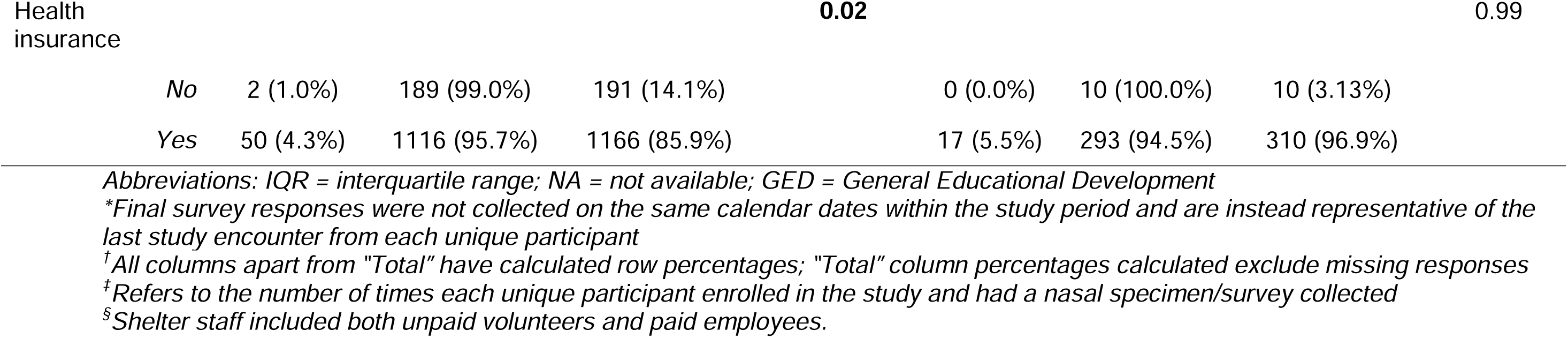
Participant characteristics by SARS-CoV-2 RT-PCR test result, by shelter staff and residents, based on last survey response*, 1 January 2020 – 31 May 2021 (N=2,930)

**Table 1b.**
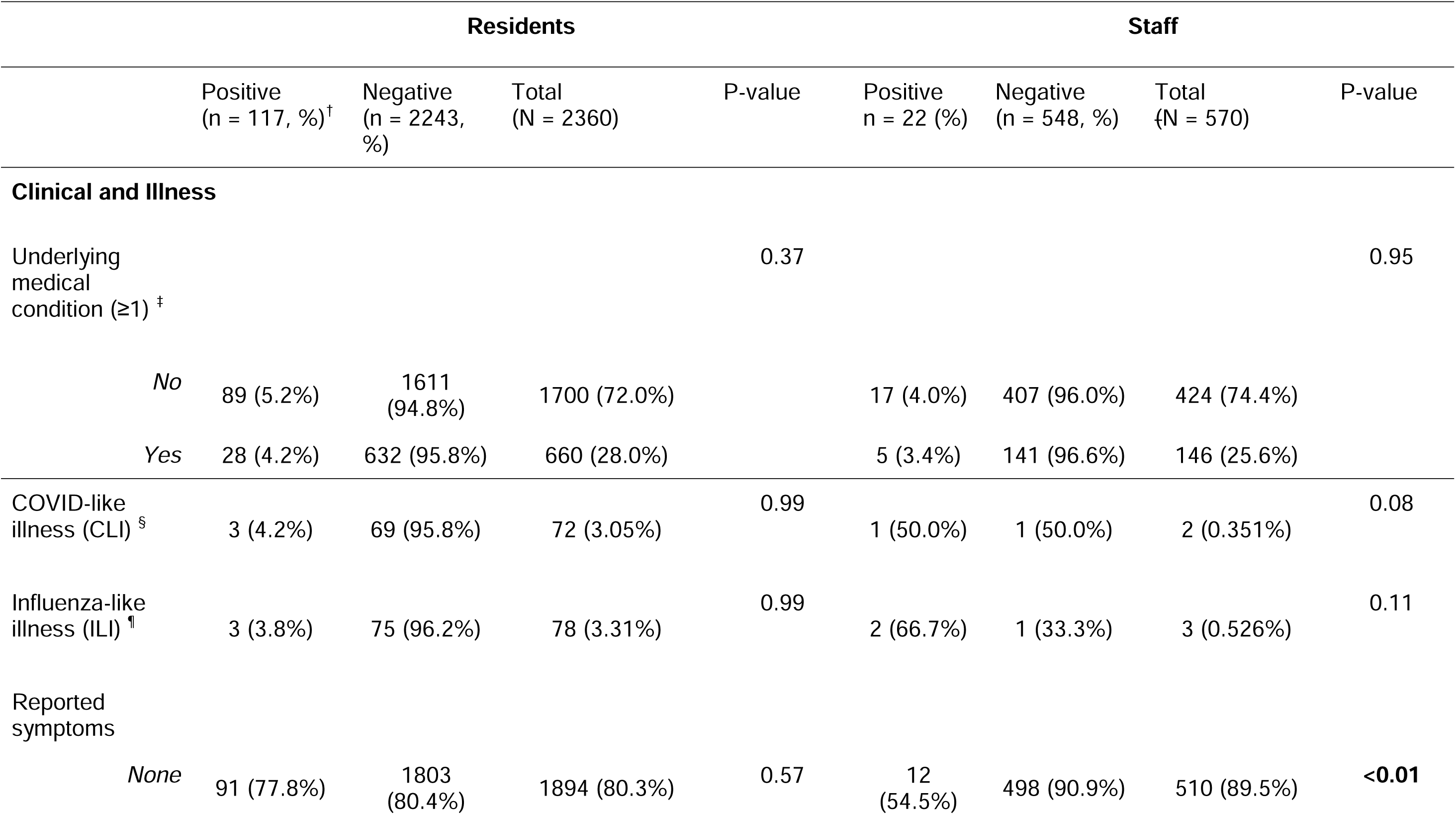
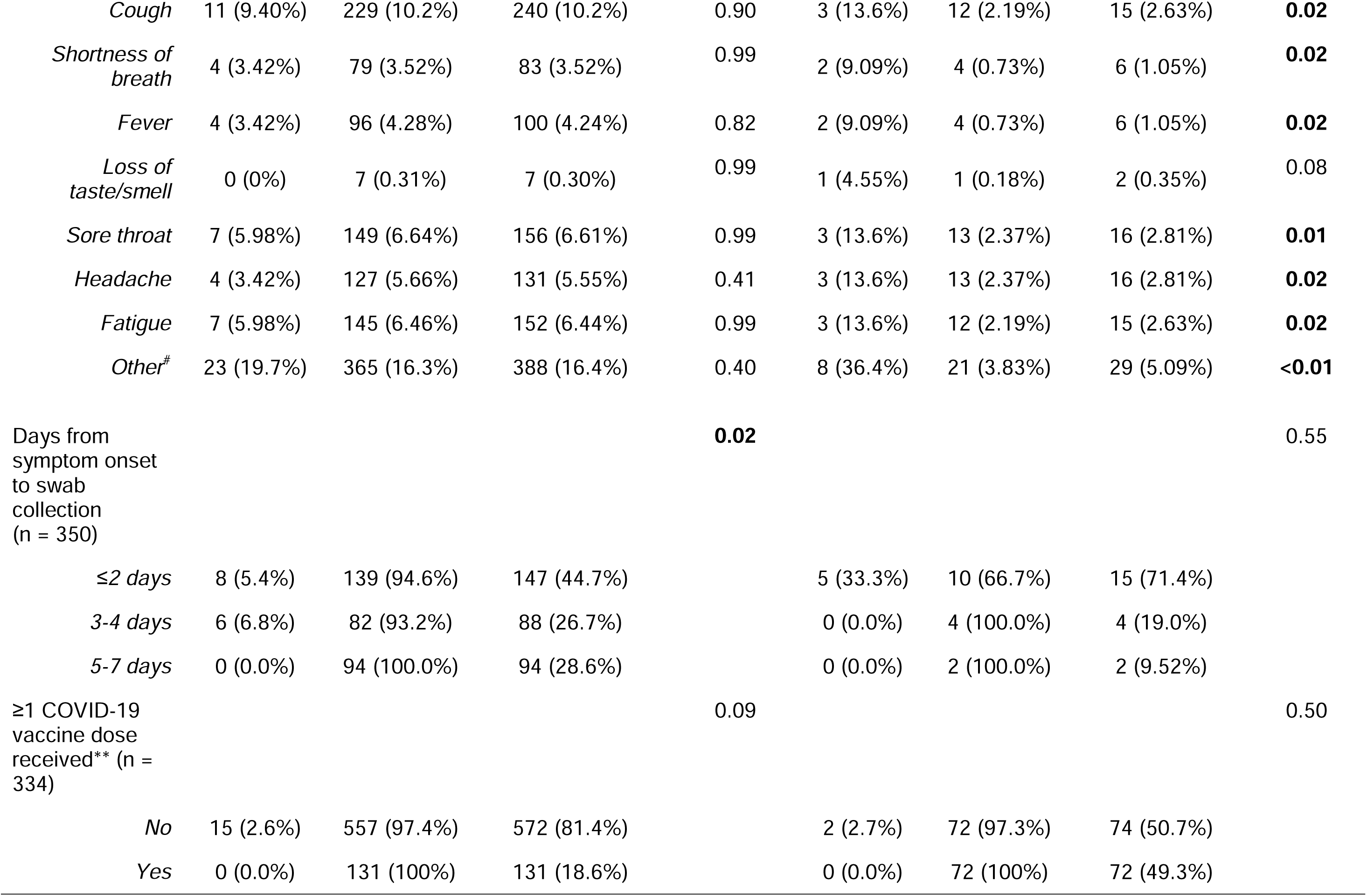
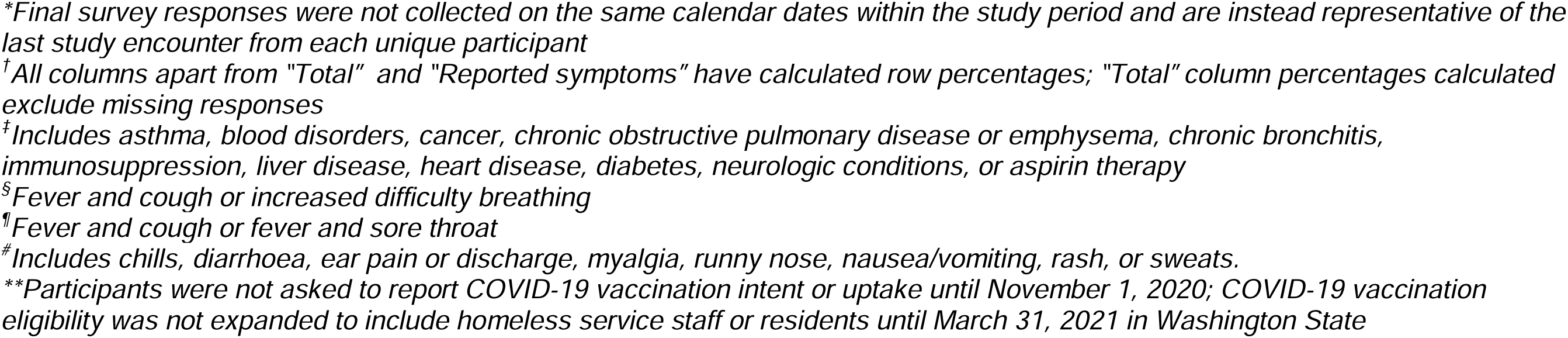
Clinical characteristics by SARS-CoV-2 RT-PCR test result, by shelter staff and residents, based on last survey response*, 1 January 2020 – 31 May 2021

| Residents |  |  |  |  | Staff |  |  |  |
| --- | --- | --- | --- | --- | --- | --- | --- | --- |
|  | Positive<br>(n = 117, %)<sup>†</sup> | Negative<br>(n = 2243,<br>%) | Total<br>(N = 2360) | P-value | Positive<br>n = 22 (%) | Negative<br>(n = 548, %) | Total<br>{N = 570) | P-value |
| Clinical and Illness |  |  |  |  |  |  |  |  |
| Underlying<br>medical<br>condition (≥1) <sup>‡</sup> |  |  |  | 0.37 |  |  |  | 0.95 |
| No | 89 (5.2%) | 1611<br>(94.8%) | 1700 (72.0%) |  | 17 (4.0%) | 407 (96.0%) | 424 (74.4%) |  |
| Yes | 28 (4.2%) | 632 (95.8%) | 660 (28.0%) |  | 5 (3.4%) | 141 (96.6%) | 146 (25.6%) |  |
| COVID-like<br>illness (CLI) <sup>§</sup> | 3 (4.2%) | 69 (95.8%) | 72 (3.05%) | 0.99 | 1 (50.0%) | 1 (50.0%) | 2 (0.351%) | 0.08 |
| Influenza-like<br>illness (ILI) <sup>¶</sup> | 3 (3.8%) | 75 (96.2%) | 78 (3.31%) | 0.99 | 2 (66.7%) | 1 (33.3%) | 3 (0.526%) | 0.11 |
| Reported<br>symptoms |  |  |  |  |  |  |  |  |
| None | 91 (77.8%) | 1803<br>(80.4%) | 1894 (80.3%) | 0.57 | 12<br>(54.5%) | 498 (90.9%) | 510 (89.5%) | <0.01 |
| *Cough* | 11 (9.40%) | 229 (10.2%) | 240 (10.2%) | 0.90 | 3 (13.6%) | 12 (2.19%) | 15 (2.63%) | **0.02** |
| *Shortness of breath* | 4 (3.42%) | 79 (3.52%) | 83 (3.52%) | 0.99 | 2 (9.09%) | 4 (0.73%) | 6 (1.05%) | **0.02** |
| *Fever* | 4 (3.42%) | 96 (4.28%) | 100 (4.24%) | 0.82 | 2 (9.09%) | 4 (0.73%) | 6 (1.05%) | **0.02** |
| *Loss of taste/smell* | 0 (0%) | 7 (0.31%) | 7 (0.30%) | 0.99 | 1 (4.55%) | 1 (0.18%) | 2 (0.35%) | 0.08 |
| *Sore throat* | 7 (5.98%) | 149 (6.64%) | 156 (6.61%) | 0.99 | 3 (13.6%) | 13 (2.37%) | 16 (2.81%) | **0.01** |
| *Headache* | 4 (3.42%) | 127 (5.66%) | 131 (5.55%) | 0.41 | 3 (13.6%) | 13 (2.37%) | 16 (2.81%) | **0.02** |
| *Fatigue* | 7 (5.98%) | 145 (6.46%) | 152 (6.44%) | 0.99 | 3 (13.6%) | 12 (2.19%) | 15 (2.63%) | **0.02** |
| *Other<sup>#</sup>* | 23 (19.7%) | 365 (16.3%) | 388 (16.4%) | 0.40 | 8 (36.4%) | 21 (3.83%) | 29 (5.09%) | **<0.01** |
| Days from symptom onset to swab collection (n = 350) |  |  |  | **0.02** |  |  |  | 0.55 |
| ≤2 days | 8 (5.4%) | 139 (94.6%) | 147 (44.7%) |  | 5 (33.3%) | 10 (66.7%) | 15 (71.4%) |  |
| 3-4 days | 6 (6.8%) | 82 (93.2%) | 88 (26.7%) |  | 0 (0.0%) | 4 (100.0%) | 4 (19.0%) |  |
| 5-7 days | 0 (0.0%) | 94 (100.0%) | 94 (28.6%) |  | 0 (0.0%) | 2 (100.0%) | 2 (9.52%) |  |
| ≥1 COVID-19 vaccine dose received** (n = 334) |  |  |  | 0.09 |  |  |  | 0.50 |
| No | 15 (2.6%) | 557 (97.4%) | 572 (81.4%) |  | 2 (2.7%) | 72 (97.3%) | 74 (50.7%) |  |
| Yes | 0 (0.0%) | 131 (100%) | 131 (18.6%) |  | 0 (0.0%) | 72 (100%) | 72 (49.3%) |  |

*\*Final survey responses were not collected on the same calendar dates within the study period and are instead representative of the last study encounter from each unique participant*
*†All columns apart from “Total” and “Reported symptoms” have calculated row percentages; “Total” column percentages calculated exclude missing responses*
*‡Includes asthma, blood disorders, cancer, chronic obstructive pulmonary disease or emphysema, chronic bronchitis, immunosuppression, liver disease, heart disease, diabetes, neurologic conditions, or aspirin therapy*
*§Fever and cough or increased difficulty breathing*
*¶Fever and cough or fever and sore throat*
*#Includes chills, diarrhoea, ear pain or discharge, myalgia, runny nose, nausea/vomiting, rash, or sweats.*
*\*\*Participants were not asked to report COVID-19 vaccination intent or uptake until November 1, 2020; COVID-19 vaccination eligibility was not expanded to include homeless service staff or residents until March 31, 2021 in Washington State*

**Table 1c.** Shelter-level characteristics by SARS-CoV-2 RT-PCR test result based on all participant encounters, 1 January 2020 – 31 May 2021 (N=12,915)

| Residents |  |  |  |  | Staff |  |  |  |
| --- | --- | --- | --- | --- | --- | --- | --- | --- |
|  | Positive<br>(n = 117, %)* | Negative<br>(n = 9729, %) | Total<br>(N = 9846) | P-value | Positive<br>(n = 22, %) | Negative<br>(n = 3047, %) | Total<br>(N = 3069) | P-value |
| Testing event |  |  |  | **<0.01** |  |  |  | **<0.01** |
| *Routine* | 87 (1.0%) | 8646 (99.0%) | 8733 (88.7%) |  | 14 (0.5%) | 2759 (99.5%) | 2773 (90.4%) |  |
| *Outbreak* | 30 (2.7%) | 1083 (97.3%) | 1113 (11.3%) |  | 8 (2.7%) | 288 (97.3%) | 296 (9.64%) |  |
| Shelter Type |  |  |  | **<0.01** |  |  |  | 0.16 |
| *Adult female* | 3 (0.7%) | 421 (99.3%) | 424 (4.31%) |  | 0 (0.0%) | 204 (100.0%) | 204 (6.65%) |  |
| *Adult male* | 33 (2.2%) | 1434 (97.8%) | 1467 (14.9%) |  | 4 (1.5%) | 260 (98.5%) | 264 (8.60%) |  |
| *Family* | 46 (1.4%) | 3337 (98.6%) | 3383 (34.4%) |  | 8 (0.7%) | 1179 (99.3%) | 1187 (38.7%) |  |
| *Mixed adult* | 32 (0.8%) | 3910 (99.2%) | 3942 (40.0%) |  | 8 (1.1%) | 750 (98.9%) | 758 (24.7%) |  |
| *Youth* | 3 (0.5%) | 627 (99.5%) | 630 (6.40%) |  | 2 (0.3%) | 654 (99.7%) | 656 (21.4%) |  |
| Sleeping arrangement |  |  |  | 0.06 |  |  |  | -- |
| *Communal* | 39 (1.3%) | 2869 (98.7%) | 2908 (29.5%) |  | NA | NA | NA |  |
| *Cubicles* | 3 (0.4%) | 791 (99.6%) | 794 (8.06%) |  | NA | NA | NA |  |
| *Private/shared family room* | 75 (1.2%) | 6069 (98.8%) | 6144 (62.4%) |  | NA | NA | NA |  |

\*All columns apart from "Total" have calculated row percentages; "Total" column percentages calculated exclude missing responses

There was a high degree of missingness for certain variables that were added mid-study as we learned more about SARS-CoV-2 and COVID-19 disease (e.g., COVID-19 vaccination status, anosmia as a self-assessed symptom). However, sensitivity analyses showed this had little effect on the associated risk factors assessed through multivariable regression.

Ethics approval was obtained from the University of Washington Human Subjects Division. The CDC determined that the study was conducted consistent with applicable federal law and CDC policy (see 45 C.F.R part 46; 21 C.F.R. part 56).

## Results

### Participant characteristics

Overall, 12,915 nasal swab specimens were collected from 2,930 unique participants from 1/1/2020 through 5/31/2020. Of these participants, 2,360 were shelter residents (80.5%) and 570 (19.5%) were shelter staff (Table 1a). Each participant was tested a median of two times (interquartile ranges (IQR) of [4] and [5] among residents and staff, respectively) over the study period. The median age of residents and staff was 37 years (range: 0–85 years) and 32.5 years (range: 18-81 years), respectively. A majority of residents self-identified as male (64.3%) compared to a majority of staff self-identifying as female (58.2%). A plurality of residents self-identified as Black (39.3%) whereas the majority of staff self-identified as White (55.0%). Receipt of seasonal influenza vaccine for the corresponding influenza season was reported by 42.5% of residents and 51.1% of staff. Among residents, 45.6% (n=611) had experienced chronic homelessness (duration ≥1 year) and 17.5% (n=191) of residents were employed.

Among unique participants, 80.3% (n=1,894, Table 1b) of residents and 89.5% (n=510) of staff were asymptomatic when specimens were collected. Among symptomatic participants (residents, n=466; staff, n=60), the most commonly reported symptoms were cough (51.5%), sore throat (33.5%) and fatigue (32.6%) for residents; cough (25.0%), fatigue (25.0%), sore throat (26.7%) and headache (26.7%) for staff. Based on their last survey response, 18.6% of residents and 49.3% of staff had received ≥1 COVID-19 vaccine dose; however, only 15% of these individuals completed their final study enrollment from 3/31/2021 onwards (when vaccine eligibility expanded to include anyone living in congregate settings)(19), limiting interpretability.

### Shelter characteristics

Table 1c presents shelter characteristics by SARS-CoV-2 test result. Nearly 90% (n=11,506) of swabs were collected during routine surveillance testing events and a plurality were collected from shelters serving mixed gender adults (36.4%, n=4,700 residents and staff). Among residents, most tests were collected from participants sleeping in private/shared rooms or rooms serving single family units (62.4%, n=6,144).

### Incidence of SARS-CoV-2 infection

A total of 139 cases of SARS-CoV-2 infection were detected over the study period. The overall estimated incidence of infection was 4.74 (95% CI 4.00-5.58) cases per 100 individuals at risk. Among unique shelter residents the incidence was 4.96 (95% CI 4.12-5.91) cases per 100 individuals at risk, compared to 3.86 (95% CI 2.43-5.79, Table 2) among staff. Incidence was highest among residents aged ≥65 years (7.69 cases per 100, 95% CI 3.90-13.35). Black participants had the highest observed incidence of infection compared to other racial groups (residents: 6.81, 95% CI 5.12-8.83; staff: 6.21, 95% CI 2.88-11.46). When stratifying by shelter type, incidence was lower at youth shelters (1.41, 95% CI 0.46-3.27) compared to adult and family shelters. Incidence was higher among symptomatic (6.84 cases per 100, 95% CI 4.84-9.35) compared to asymptomatic individuals (4.28, 95% CI 3.51-5.17, Figure 1).

**Figure 1.**
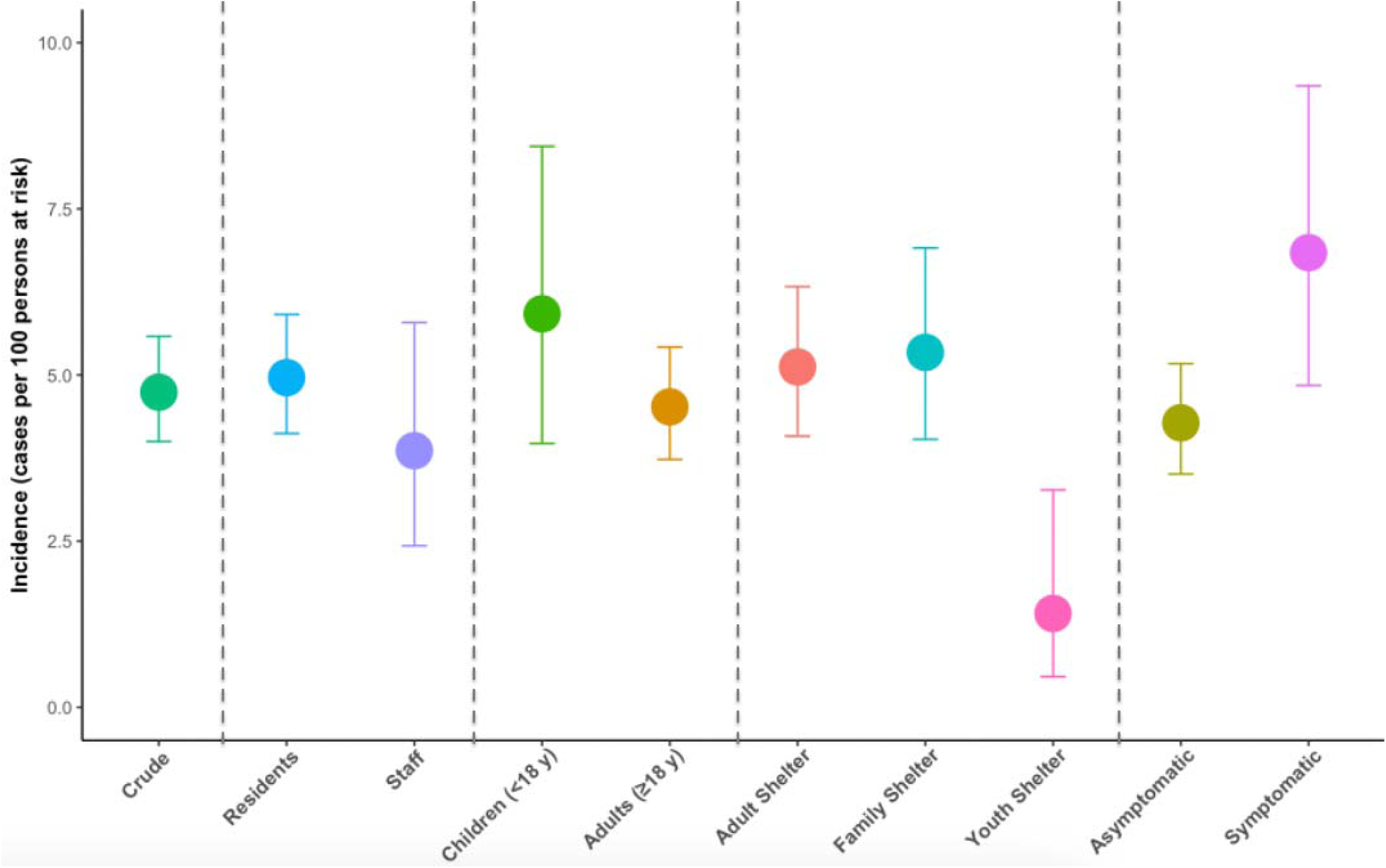
Crude incidence estimates among all unique participants, plus stratifications: (a) resident vs. staff; (b) children vs. adults; (c) shelter type (adult, family, youth); (d) asymptomatic vs. symptomatic (≥1 symptom)

**Table 2.** Incidence estimates for SARS-CoV-2 RT-PCR positive test result among unique study participants. Characteristics are based on last survey response*

| Characteristic | Residents |  |  | Staff |  |  |
| --- | --- | --- | --- | --- | --- | --- |
|  | Positive | Total tested | Incidence (95% CI) per 100 persons at risk | Positive | Total tested | Incidence (95% CI) per 100 persons at risk |
| OVERALL | 117 | 2360 | 4.96 (4.12 – 5.91) | 22 | 570 | 3.86 (2.43 – 5.79) |
| Age group |  |  |  |  |  |  |
| <5 y | 9 | 174 | 5.17 (2.39 – 9.59) | -- | -- | -- |
| 5-11 y | 13 | 193 | 6.74 (3.63 – 11.24) | -- | -- | -- |
| 12-17 y | 6 | 106 | 5.66 (2.11 – 11.91) | -- | -- | -- |
| 18-49 y | 45 | 1140 | 3.94 (2.89 – 5.25) | 18 | 452 | 3.98 (2.38 – 6.22) |
| 50-64 y | 33 | 604 | 5.46 (3.79 – 7.59) | 4 | 98 | 4.08 (1.12 – 10.12) |
| ≥65 y | 11 | 143 | 7.69 (3.90 – 13.35) | 0 | 20 | -- |
| Sex |  |  |  |  |  |  |
| Female | 35 | 824 | 4.25 (2.98 – 5.86) | 16 | 325 | 5.92 (2.84 – 7.87) |
| Male | 79 | 1484 | 5.32 (4.24 – 6.59) | 6 | 233 | 2.58 (0.95 – 5.52) |
| Race |  |  |  |  |  |  |
| American Indian and Alaskan Native | 4 | 91 | 4.40 (1.21 – 10.87) | 0 | 7 | -- |
| *Asian* | 0 | 59 | -- | 1 | 48 | 2.08 (0.05 – 11.07) |
| *Black/African American* | 52 | 764 | 6.81 (5.12 – 8.83) | 9 | 145 | 6.21 (2.88 – 11.46) |
| *Multiracial* | 9 | 202 | 4.46 (2.06 – 8.29) | 1 | 22 | 4.55 (0.12 – 22.84) |
| *Native Hawaiian and Other Pacific Islander* | 2 | 124 | 1.61 (0.20 – 5.70) | 4 | 14 | 28.57 (8.39 – 58.10) |
| *White* | 27 | 705 | 3.83 (2.54 – 5.52) | 6 | 288 | 2.08 (0.77 – 4.48) |
| Hispanic/Latinx |  |  |  |  |  |  |
| Yes | 17 | 399 | 4.26 (2.50 – 6.73) | 2 | 55 | 3.63 (0.44 – 12.53) |
| No | 96 | 1906 | 5.03 (4.09 – 6.12) | 20 | 511 | 3.91 (2.41 – 5.98) |
| Shelter type |  |  |  |  |  |  |
| Adult | 68 | 1329 | 5.12 (3.99 – 6.44) | 12 | 235 | 5.11 (2.67 – 8.75) |
| Family | 46 | 851 | 5.41 (3.98 – 7.14) | 8 | 161 | 4.97 (2.17 – 9.56) |
| Youth | 3 | 180 | 1.67 (0.35 – 4.79) | 2 | 174 | 1.15 (0.14 – 4.09) |

*\*Final survey responses were not collected on the same calendar dates within the study period and are instead representative of the last study encounter from each unique participant*

Among 2,930 persons tested, SARS-CoV-2 infections peaked in Week 37 (9/6/2020-9/12/2020) of 2020 with 15 unique participants testing positive, with additional peaks in infections observed in Week 17 (4/19/2020-4/25/2020) and Week 51 (12/13/2020-12/19/2020) of 2020 and continued detection observed through the duration of the study period (Figure 2a). Among 12,915 tests performed, SARS-CoV-2 test positivity peaked earlier at 9% in epidemiologic week 17 of 2020 (Figure 2b). The proportion of participant encounters self-reporting at least one dose of a COVID-19 vaccine is represented in Figure 2c; we observed a consistent trend towards increased vaccine uptake from Week 4 (1/24/2021-1/30/2021) in 2021 through the end of the study period.

**Figure 2a-c.**
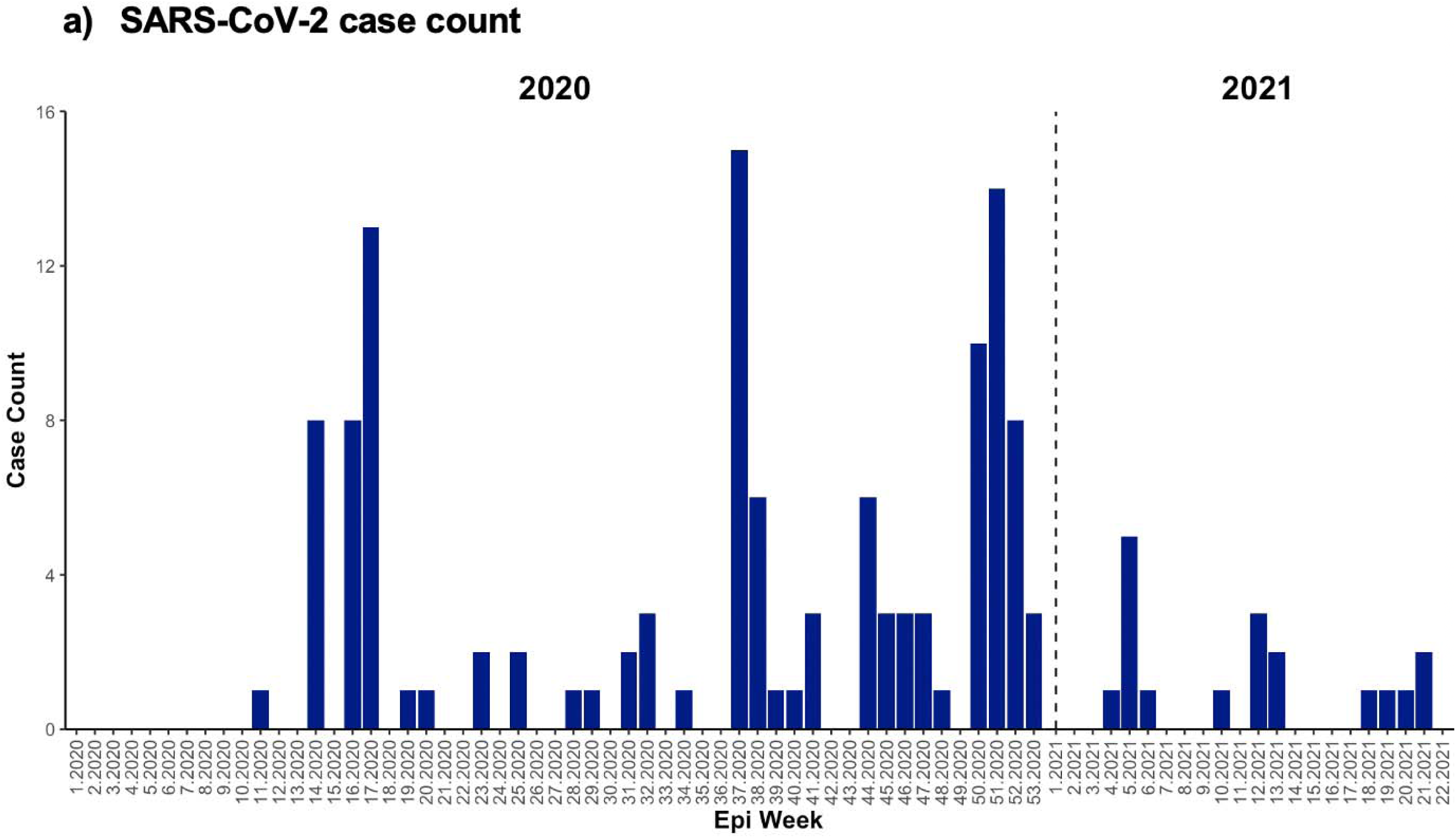
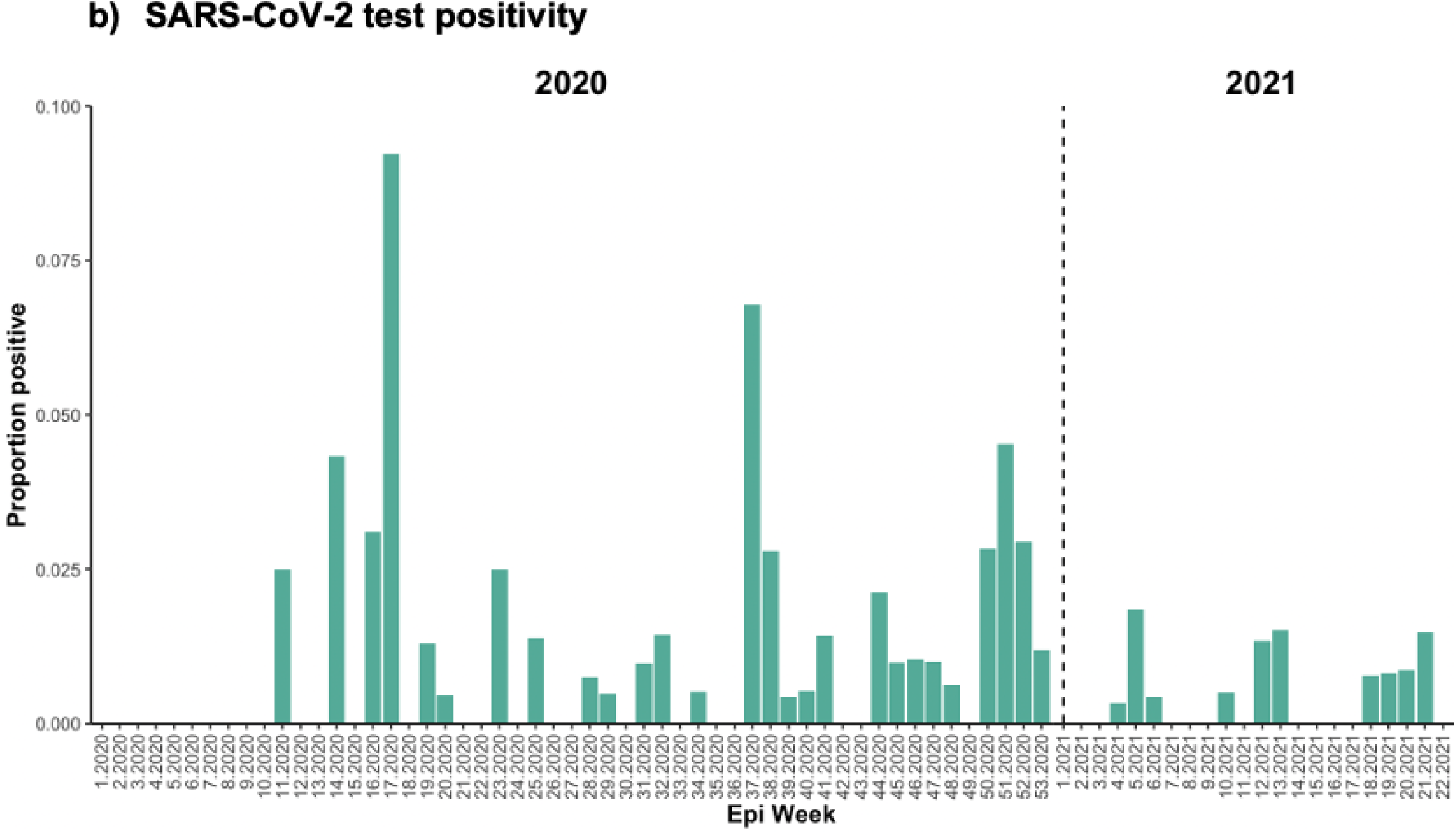
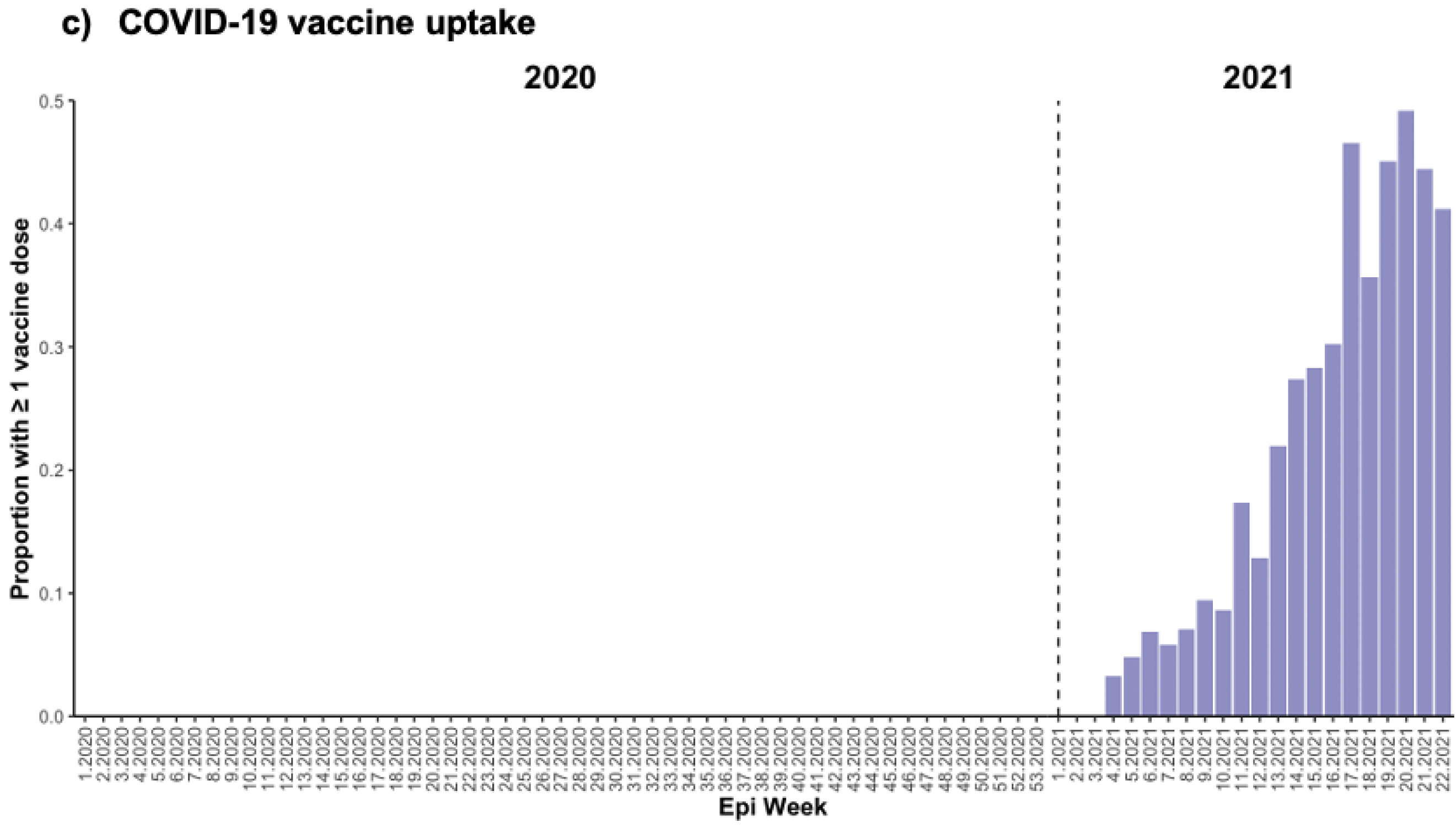
Epidemic Curves of SARS-CoV-2 case count (a; N=139); test positivity (b; N=139/12,915); and COVID-19 vaccine uptake (≥1 dose) (c; N=597/12,915) by Epidemiological Week

Combining data from residents and staff, most infections were asymptomatic at time of detection (74%, 103/139, Table 1b) and detected during routine surveillance (73%, 101/139, Table 1c). Overall test positivity was 1.2%, however, outbreak testing yielded higher positivity (2.7%, 38/1,409 vs. 0.9%, 101/11,506, Table1c).

### Factors associated with SARS-CoV-2 infection

Based on unique participants’ last surveys (N=2,930), unadjusted models show that among residents, Black race (OR=1.83, 95% CI 1.15-2.99) was significantly associated with higher odds of SARS-CoV-2 infection, whereas residents who were current smokers had a decreased odds of infection (OR=0.53, 95% CI 0.36–0.79). Adjusting for other model variables (Table 3a), residents who smoked had 66% (aOR=0.34, 95% CI 0.20–0.59) lower odds of SARS-CoV-2 infection compared with non-smokers, and residents who had received that season’s influenza vaccine had 46% (aOR=0.54, 95% CI 0.33–0.90) lower odds of infection compared with those who had not received an influenza vaccine. Among staff, Native Hawaiian and Other Pacific Islander (NHPI) race was also identified with a significant association, however the validity of this finding is undermined by the small sample size of NHPI participants.

**Table 3a.**
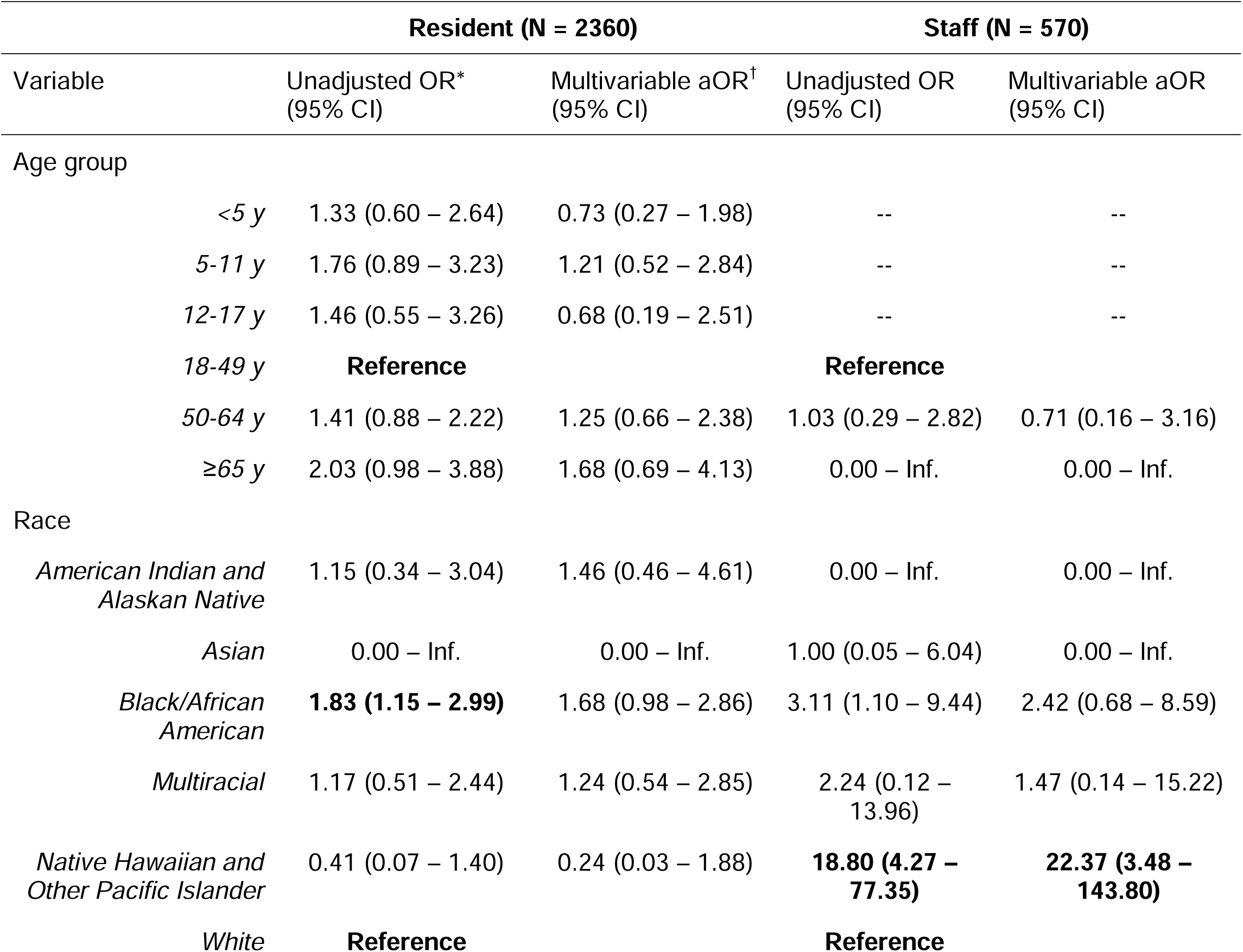
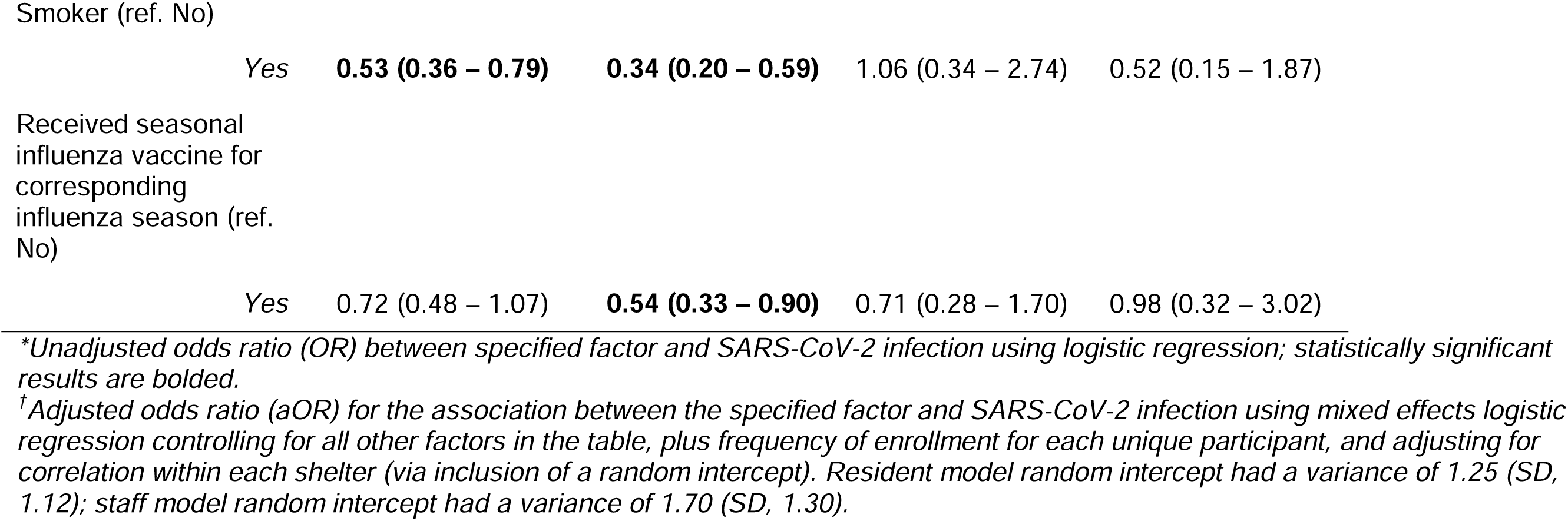
Results of logistic regression analyses, unadjusted and adjusted, for factors associated with SARS-CoV-2 infection among residents and among staff, regardless of symptom profile, 1 January 2020 – 31 May 2021

**Table 3b.**
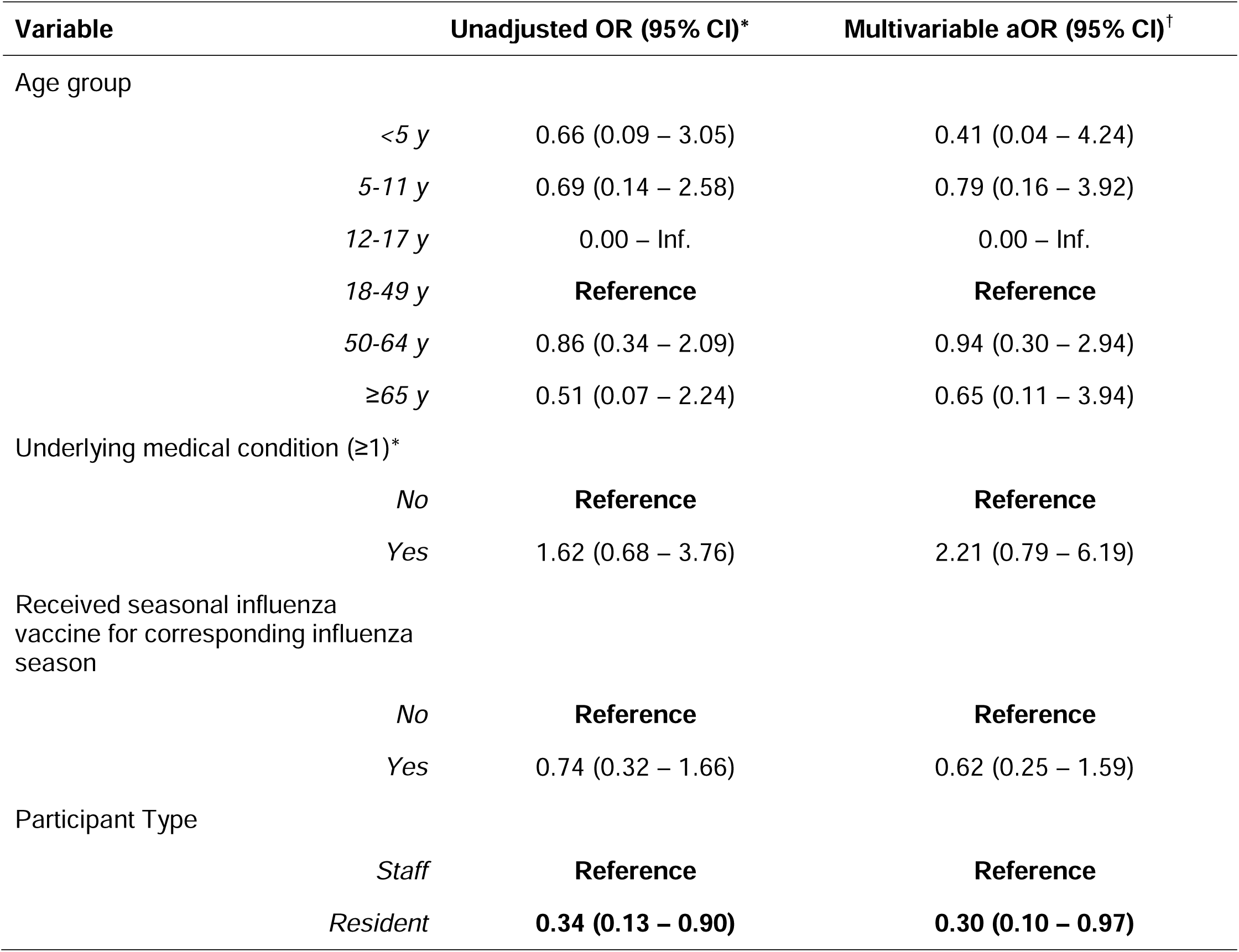
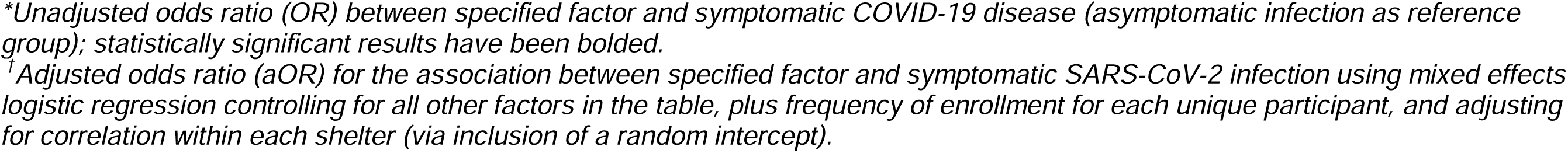
Factors associated with symptomatic COVID-19 disease (n=36) among all unique participants with a SARS-CoV-2 infection detected (N=139)

| Variable | Unadjusted OR (95% CI)* | Multivariable aOR (95% CI)<sup>†</sup> |
| --- | --- | --- |
| Age group |  |  |
| <5 y | 0.66 (0.09 – 3.05) | 0.41 (0.04 – 4.24) |
| 5-11 y | 0.69 (0.14 – 2.58) | 0.79 (0.16 – 3.92) |
| 12-17 y | 0.00 – Inf. | 0.00 – Inf. |
| 18-49 y | **Reference** | **Reference** |
| 50-64 y | 0.86 (0.34 – 2.09) | 0.94 (0.30 – 2.94) |
| ≥65 y | 0.51 (0.07 – 2.24) | 0.65 (0.11 – 3.94) |
| Underlying medical condition (≥1)* |  |  |
| No | **Reference** | **Reference** |
| Yes | 1.62 (0.68 – 3.76) | 2.21 (0.79 – 6.19) |
| Received seasonal influenza vaccine for corresponding influenza season |  |  |
| No | **Reference** | **Reference** |
| Yes | 0.74 (0.32 – 1.66) | 0.62 (0.25 – 1.59) |
| Participant Type |  |  |
| Staff | **Reference** | **Reference** |
| Resident | **0.34 (0.13 – 0.90)** | **0.30 (0.10 – 0.97)** |

*\*Unadjusted odds ratio (OR) between specified factor and symptomatic COVID-19 disease (asymptomatic infection as reference group); statistically significant results have been bolded.*
*†Adjusted odds ratio (aOR) for the association between specified factor and symptomatic SARS-CoV-2 infection using mixed effects logistic regression controlling for all other factors in the table, plus frequency of enrollment for each unique participant, and adjusting for correlation within each shelter (via inclusion of a random intercept).*

### Individual factors associated with symptomatic COVID-19 disease

When assessing factors associated with symptomatic COVID-19 among unique participants (n=36) with SARS-CoV-2 infections detected (n=139), the only variable significantly associated with symptomatic infection was staff versus resident status (Table 3b). Adjusting for other variables, shelter residents had 70% (0.30, 95% CI 0.10–0.97) lower odds of reporting ≥1 new or worsening symptom within 7 days of sample collection compared to shelter staff.

## Discussion

From 1/1/2020 through 5/31/2021, we conducted active surveillance in 23 homeless shelters in King County, Washington. We detected an incidence of 4.74 SARS-CoV-2 cases per 100 persons at risk and identified risk factors associated with infection. Most infections were detected during routine surveillance and staff were more likely to report symptoms than residents among those infected.

Among King County’s estimated population of 2.26 million people, there were 106,347 confirmed cases reported from the start of the pandemic through 5/31/21; an incidence of 4.71 cases per 100 persons can be calculated based on these figures.(20) This striking similarity in disease burden when compared to our findings may be reflective of our study’s early focus on testing asymptomatic individuals: if only symptomatic individuals in our study received testing, as was largely the case for the greater King County community during the early months of the pandemic, the denominator of persons-at-risk may have been smaller but more likely to test positive. Calculated incidence as a result would have been higher in the shelters compared to the general population. As of 1/4/22, PEH comprised 1.4% of COVID-19 cases in King County but only represented 0.5% of its population.(21) Additionally, 12.4% of PEH cases reported to PHSKC were hospitalized due to COVID-19 disease compared to 3.3% in King County’s general population.(21) A population level study in Wales, UK found that SARS-CoV-2 prevalence among PEH was lower than the general population.(22) However, this study and others may not account for the differential healthcare seeking behavior or time at risk between

PEH and non-PEH which may result in an under-detection of infections when testing is performed in a clinical setting. Despite differential testing methodologies between the general population and our study, we did observe similar temporal trends and spikes in test positivity in mid-April and late-December 2020, supporting previously published evidence of genetic relationships and synergistic transmission dynamics with the broader community.(20,23)

A model of SARS-CoV-2’s potential effect among the U.S.’s PEH population published in late March 2020 projected that 40% would be infected at pandemic’s peak due to conditions at homeless service sites and a high prevalence of medical comorbidities.(24) Comparable cross-sectional results were reported in an adult shelter in Boston, MA, USA where an outbreak investigation yielded 36% test positivity, while one in San Francisco, CA, USA yielded even higher test positivity (67%)(8,25) These estimates and results from specific outbreak testing demonstrate a substantially higher burden than what was observed in this study during similar time periods, likely due to discrepant testing methodologies (e.g., data from the San Francisco and Boston studies were collected from contact tracing efforts post-outbreak identification), in addition to regional differences in background community prevalence. A systematic review of studies addressing COVID-19 and health-related outcomes in PEH and shelters’ staff estimated a pooled SARS-CoV-2 prevalence of 32% among PEH in an outbreak context compared to 2% in the absence of an acute outbreak.(5) This analysis, however, was limited by the relatively short observational periods of its studies.

A substantial proportion of the SARS-CoV-2 infections in our study were asymptomatic at the time specimen were collected. Prior studies of seropositive cases in shelters found that 68-85% of all cases had no symptoms at time of testing.(9,26–28) An Atlanta, Georgia, USA, study of symptom evolution of PEH staying in isolation hotels after testing positive for SARS-CoV-2 found 32% of community referrals became symptomatic after testing positive.(29) Our participants were not longitudinally followed after detection, but our data add to the evidence that asymptomatic routine testing of all staff and residents is important in congregate living settings.

SARS-CoV-2-positive staff were more likely to report symptoms than residents. This has important implications. First, residents might be hesitant or unable to report symptoms.(30) Second, regardless of policies in place, staff may have worked while experiencing COVID-19 symptoms due to unavailability of paid sick leave, fear of job loss,(31) or dedication to their roles as essential workers. A study of SARS-CoV-2 molecular epidemiology in shelters found evidence that most infections were the result of intra-shelter transmission while staff working across multiple facilities may have introduced the virus into some of the facilities.(23) This finding may also be an artifact of surveillance timing given that residents were less consistently surveilled than staff and therefore their positive tests may have been well into their course of infection (i.e. persistent positives).

We found that the highest test positivity was detected in adult male shelters, all of which provided services 24 hours per day. Comparatively, the youth shelters included in this study, which had the lowest observed test positivity, closed services during the day, likely reducing social mixing in both formal and informal communal spaces. King County’s swift creation of nearly 2,000 new spaces (i.e. beds, isolation or quarantine areas) in homeless service sites likely had a substantial impact on mitigating transmission.(29,32,33) Specifically, protocols enacted by PHSKC that relocated consenting SARS-CoV-2-positive shelter residents from our study sites to medically-attended isolation and quarantine units likely reduced incidence. The lack of significant association between sleeping arrangements and risk of infection in our study suggests that other factors, such as intra-shelter social mixing patterns, are facilitating virus transmission, especially in facilities with non-congregate sleeping arrangements but shared hygiene and communal spaces. The provision of high-efficiency particulate air (HEPA) filters distributed by PHSKC to shelters during the pandemic may have also reduced particulate matter exposure, and subsequently impacted SARS-CoV-2 incidence, in our study population.(34) However, a simulation study found that in shelters at high risk of a SARS-CoV-2 outbreak, no additional non-pharmaceutical infection control strategy is likely to prevent outbreaks.(35) This evidence supports the prioritization of non-congregate housing options for PEH.

Our findings suggest that, over a prolonged surveillance period, environmental and behavioral factors may obfuscate associations between SARS-CoV-2 infection and individual-level risk factors. The protective association observed between influenza vaccination and SARS-CoV-2 infection is consistent with published literature(36–38), as well as the negative association between smoking and infection.(39) However these studies were subject to methodological limitations and probable confounding variables,(39–42) and there is no consensus about either relationship. Furthermore, these associations were only observed among residents, limiting our ability to conclude if reflective of true biological mechanisms, behavioral differences, or unobserved confounding variables.

This study is subject to several limitations. The repeated cross-sectional nature of this study in an open population where participant time-at-risk was not calculable likely resulted in an underestimation of the true disease burden. For these reasons, more specific measures of disease occurrence such as “cumulative incidence” or “incidence rate” could not be applied to this study population. Another limitation is our inability to differentiate between pre-symptomatic, asymptomatic, and convalescent cases due to the cross-sectional design of this study and limiting self-report of new or worsening from <7 days. We also do not have complete infection history for study participants prior to entering the shelters, likely resulting in an underestimation of incident infection – especially among residents who are less consistently surveilled in this study population – given the protective effect of SARS-CoV-2 infection history.(43) Finally, organizational infection prevention methods instituted to mitigate transmission were not routinely collected in this study and therefore their impact could not be examined.

## Conclusion

To our knowledge, this is one of the first studies to capture temporal trends and estimate incident SARS-CoV-2 infections among shelter populations through prolonged, active surveillance efforts. Our findings suggest that routine surveillance for SARS-CoV-2 that includes testing of all persons, regardless of symptoms, is essential in ascertaining the true burden of disease among residents and staff of congregate settings. As the COVID-19 pandemic evolves,(44) additional studies are recommended to assess the cost-effectiveness of routine shelter-based SARS-CoV-2 testing and impact of transmission mitigation efforts in low resource, congregate living settings.

## Disclaimers

The findings and conclusions in this report are those of the authors and do not necessarily represent the view of the US Centers for Disease Control and Prevention.

## Data Availability

All data produced in the present study are available upon reasonable request to the authors.

## Acknowledgements

We thank the shelter staff and program managers for their cooperation and collaboration throughout the participant recruitment process. A special thanks to all participants and the research assistants who assisted with data collection.

