## Supplementary Material for "Incidence of SARS-CoV-2 infection and associated risk factors among staff and residents at homeless shelters in King County, Washington: an active surveillance study"

Statistical Analysis

Encounters from the same participant were linked and assigned a unique identifier *post hoc* using their name and DOB. Incongruous name spellings due to clerical error were addressed using a function of the Levenshtein distance, a metric used to measure the differences between two character strings. Survey records were manually assigned to the same individual if the two names fell above a pre-specified value of similarity (>0.8 in the interval [0,1]) and had the same DOB. If two survey records had the same name but one-digit discrepancy in the DOB, the same unique identifier was assigned.

While it was not possible to determine person-time at risk for this population, on average resident participants self-reported a 5-month duration of stay at the shelter where their sample was collected. Anecdotal evidence provided by management suggests shelter staff were employed on average 8-12 months over the study period. Our assumption that most infections were captured in this study is reasonable as outbreak testing events were also initiated by positive test results detected outside of the study’s surveillance as a result of our close collaboration with public health and shelter management. We also collected survey data on past positive SARS-CoV-2 test results from 11/1/2020 - 5/31/2021 and found that the aggregate number of infections detected by our surveillance closely matched those self-reported by participants over this time period.

Specimen Testing Methods

Nasal swabs were transported to the University of Washington laboratory in Universal Viral Transport Medium (Becton Dickinson, Franklin, NJ) in ice-packed coolers and stored at 4˚C prior to testing. Testing was performed at the Brotman Baty Institute for Precision Medicine. Total nucleic acids were extracted (MagnaPure, Roche) and tested for the presence of 27 respiratory pathogens by TaqMan reverse-transcription polymerase chain reaction (RT-PCR) on the OpenArray platform (ThermoFisher), and for SARS-CoV-2 using a laboratory-developed test or research assay. For the laboratory-developed test, SARS-CoV-2 detection was performed using RT-PCR with probe sets targeting Orf1b and S with FAM fluor (Life Technologies 4332079 assays # APGZJKF and APXGVC4APX) multiplexed with an RNaseP probe set with VIC or HEX fluor (Life Technologies A30064 or IDT custome) each in duplicate on a QuantStudio 6 instrument (Applied Biosystems). The research assay employs only the Orf1b and RNaseP multiplexed RT-PCR in duplicate.

Shelter specimens collected from 2/25/2020 until 3/9/2020 were tested for SARS-CoV-2 using the research assay in real time. Specimens collected after 3/9/2020 were tested for SARS-CoV-2 using the laboratory-developed test under an Emergency Use Authorization issued by Washington State. Specimens collected prior to 2/25/2020 were tested retrospectively using a single replicate RT-PCR research assay to detect SARS-CoV-2 Orf1b.

Figure S1 – Timeline of testing methodology changes*

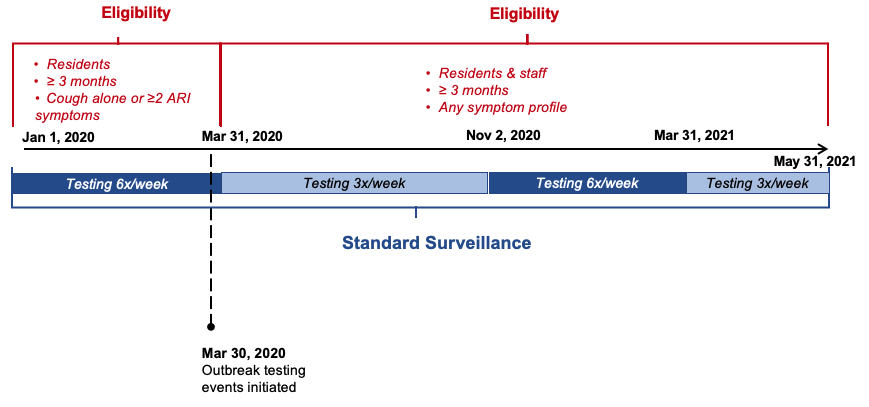

* *ARI symptoms include: fever/feverishness, cough, sore throat, shortness of breath, myalgia, headache, rhinorrhea, anosmia, nausea/vomiting; plus diarrhoea, rash and ear pain/ear discharge in participants < 18 years.*

Figure S2 – Study flow

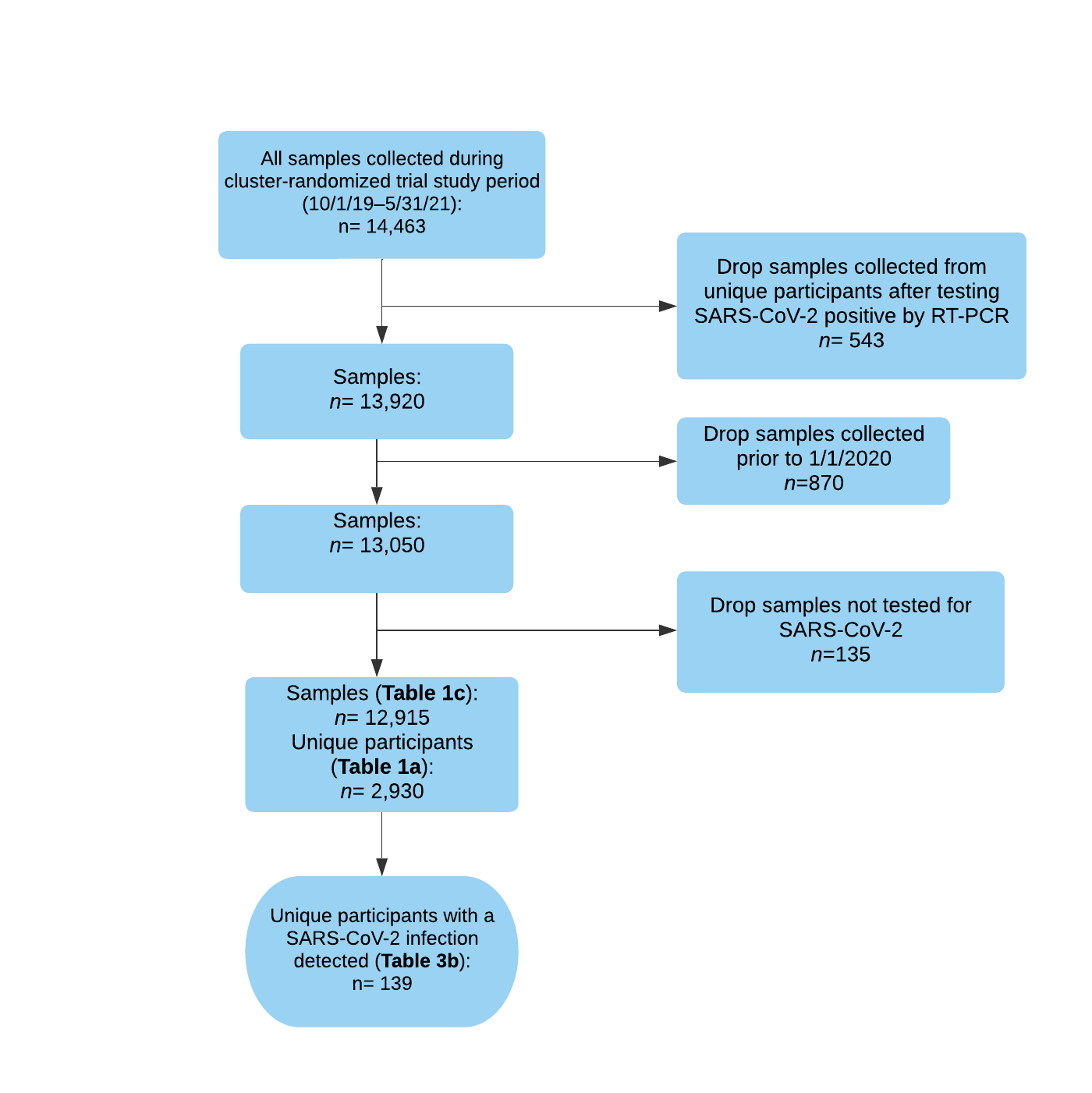

Table S1 – All shelter sites where sample collection occurred, routine surveillance and outbreak testing, October 1, 2019 – May 31, 2021

| **Shelter** | | **Max. capacity** | | **Resident sex** | | **Resident age range** | | **Sleeping arrangements available** | |
| --- | --- | --- | --- | --- | --- | --- | --- | --- | --- |
| ***Routine surveillance sites*** | | | | | | | | | |
| A | | 60 | | Female | | ≥ 18 years | | Communal bunk beds | |
| B | | 100 | | Mixed | | ≥ 18 years | | Communal bunk beds | |
| C | | 45 | | Mixed | | 18 - 25 years | | Communal floor mats and bunks beds | |
| D | | 185 | | Mixed | | All ages (family shelter) | | Private rooms / shared rooms / communal floor mats | |
| E | | 70 | | Mixed | | All ages (family shelter) | | Private rooms / shared rooms / communal floor mats | |
| F | | 60 | | Male | | ≥ 18 years | | Communal bunk beds | |
| G* | | 275 | | Mixed | | ≥ 18 years | | Private rooms / shared rooms | |
| H* | | 275 | | Mixed | | All ages (family shelter) | | Private rooms / shared rooms | |
| I* | | 45 | | Male | | ≥ 50 years | | 5 person dorms | |
| J* | | 34 | | Male | | ≥ 18 years | | Individual open cubicles | |
| K* | | 75 | | Mixed | | ≥ 18 years | | Individual open cubicles | |
| L | | 200 | | Mixed | | ≥ 18 years | | Communal bunk beds | |
| M | | 212 | | Male | | ≥ 50 years | | Communal floor mats | |
| N* | | 46 private rooms | | Mixed | | All ages (family shelter) | | Private rooms / shared rooms | |
| O | | 100 | | Mixed | | All ages (family shelter) | | Private rooms / shared rooms / communal floor mats | |
|  | Shelter | | Max. capacity | | Resident sex | | Resident age range | | Sleeping arrangements available |
|  | ***Outbreak testing sites*** | | | | | | | | |
|  | other_A*^†^* | | 100 | | Male | | ≥ 50 years | | Communal floor mats |
|  | other_B | | 100 | | Mixed | | ≥ 18 years | | Private apartments |
|  | other_C | | 150 | | Mixed | | ≥ 18 years | | Communal floor mats |
|  | other_D | | 234 | | Mixed | | ≥ 18 years | | Private apartments |
|  | other_E | | 49 | | Male | | ≥ 50 years | | Communal floor mats |
|  | other_F | | 50 | | Mixed | | All ages (family shelter) | | Private rooms / shared rooms / communal floor mats |
|  | other_G | | 18 | | Mixed | | <18 years | | Communal bunk beds |
|  | other_H | | 20 | | Mixed | | 18 - 25 years | | Communal bunk beds |

** Shelter facilities where residents and staff were relocated to enable improved adherence to COVID-19 infection and prevention control measures during the COVID-19 pandemic.*

*^†^ Shelters “other_A” through “other_H” used as naming mechanism for sites where only intermittent outbreak testing was conducted*
